# Helmet Use Among E-Bike, Pedal Bike, and E-Scooter Riders in Canberra: Retrospective Data Analysis of Head Injury Presentations (Phase 3) [Protocol]

**DOI:** 10.64898/2026.03.04.26347649

**Authors:** Alan Silburn

**Author notes:** Correspondence, Tele: 0449 107 944.

## Abstract

**Background:** Helmet use is a proven safety measure that reduces the risk of head injury among cyclists and e-scooter riders. Despite legal requirements for pedal bikes and e-bikes in Australia, compliance varies, particularly among users of electric vehicles. The growing popularity of e-bikes and e-scooters in urban areas presents new public health challenges, yet observational data on helmet use, behavioural determinants, and the effectiveness of safety interventions remain limited.

**Aim:** Phase 3 of the Helmet Use in Canberra study aims to characterise head injury presentations associated with cycling and e-scooter use and examine their association with helmet use and injury severity.

**Methods:** De-identified emergency department records from The Canberra Hospital will be retrospectively analysed for presentations involving cycling or e-scooter-related head injuries during the study period. Extracted variables will include age, sex, vehicle type, documented helmet use, injury diagnosis, severity indicators, and date/time of presentation. Descriptive statistics will summarise injury patterns, while regression analyses will evaluate associations between helmet use and injury severity, controlling for demographic and contextual factors. Sensitivity analyses will address missing helmet data and subgroup differences by vehicle type, age, and gender.

**Expected Results:** It is hypothesised that lower helmet use will correlate with higher rates and greater severity of head injury presentations. Findings will provide a population-level perspective on helmet effectiveness, inform local injury prevention strategies, and guide public safety interventions.

**Trial Registration:** Australian and New Zealand Clinical Trials Registry (ANZCTR) [ACTRN12626000245392]

## 1. SYNOPSIS

Helmet use is a well-established safety measure that significantly reduces the risk of head injury among cyclists and e-scooter riders. Despite Australian legislation mandating helmet use for pedal bikes and e-bikes, compliance remains variable, particularly among users of electric vehicles. The increasing popularity of e-bikes and e-scooters in urban environments presents new challenges for injury prevention. Understanding helmet-wearing behaviour, evaluating the impact of targeted interventions, and linking observational findings to hospital presentations are critical for informing evidence-based public safety policies.

This sub-study forms part of a larger study that employs a multi-method approach comprising three complementary components: a quasi-experimental observational study of helmet use in public settings, retrospective analysis of head injury presentations at The Canberra Hospital, and a cross-sectional survey assessing public attitudes toward helmet use and deterrent fines. Observational data will be collected across three high-traffic urban bike paths, with pre- and post-installation of signage emphasising either health benefits or legal penalties. Hospital data will provide context regarding injury outcomes associated with helmet use trends. Survey data will inform potential policy strategies to increase compliance.

The primary objective of the larger study is to assess helmet use rates among riders of e-bikes, pedal bikes, and e-scooters and to evaluate the effectiveness of signage interventions. Secondary objectives include examining demographic differences in helmet use, exploring associations with head injury presentations, and assessing public perceptions of deterrent fines. This protocol outlines study rationale, design, methodology, ethical considerations, and anticipated outcomes to ensure robust and meaningful findings for injury prevention in Canberra.

## 2. RATIONALE / BACKGROUND

Head injuries associated with cycling and micromobility vehicles represent a substantial public health concern, with consequences ranging from mild concussions to severe traumatic brain injury. Helmet use consistently mitigates the risk of head injury, with protective effects well documented across conventional bicycles, e-bikes, and e-scooters (1,2). For example, a meta-analysis of over 64,000 cyclist crash cases estimated that helmet use is associated with ∼51% lower risk of head injury and ∼69% lower risk of serious head injury compared with non-use, supporting widespread helmet use in cycling safety planning (1). Moreover, Høye’s review suggests that mandatory helmet legislation is associated with reductions of ∼20% in head injuries and ∼55% in serious head injuries (3). In the Australian context, the introduction of universal bicycle helmet laws in the early 1990s was followed by an immediate ∼46% decline in cycling fatality rates, though the effect on nonfatal head injuries is less consistently documented (4).

However, compliance remains inconsistent, particularly among users of electric mobility devices in urban areas. Observational studies in Australia show significant proportions of e-scooter riders not wearing helmets even where mandated (5). A study in Melbourne found that ∼50% of e-scooter injury presentations involved head injuries, while only one-third of riders reported wearing helmets (6). The rapid growth in e-scooter adoption has been mirrored by increasing hospital presentations for related injuries and calls for updated regulatory frameworks (7,8), highlighting the need to explore factors influencing helmet-wearing behaviour.

Previous observational and epidemiological studies suggest that behavioural, infrastructural, and policy factors influence helmet-wearing behaviour. Interventions such as signage emphasising health benefits or legal penalties may promote compliance, though evidence from Australian urban settings is still limited. Complementing observational data with hospital head injury records enables a triangulated view of how helmet behaviour maps to clinical outcomes, while survey data can shed light on the acceptability of enforcement strategies and what might drive behavioural change. In addition, legal context and enforcement levels may further shape compliance patterns.

Helmet laws and fines vary widely across Australian jurisdictions, which may influence helmet-wearing behaviour. In the ACT, riders face a fine of AUD 121 for not wearing a helmet, comparatively modest next to AUD 344 in New South Wales and Tasmania, AUD 227 in Victoria, and AUD 205 in South Australia. Fines are lower in Queensland (AUD 137), Western Australia (AUD 50), and the Northern Territory (AUD 25) (9). Enforcement intensity also differs, with NSW police reportedly issuing ∼500 fines per month (9). These variations suggest that both fine magnitude and policing practices may shape helmet-wearing behaviour, providing critical context for interpreting compliance trends in Canberra and for designing targeted interventions such as signage emphasising health benefits or legal penalties.

Thus, linking helmet use behaviour, signage-based interventions, and hospital head injury trends offers a promising approach to inform effective local injury prevention policy in Canberra.

## 3. AIMS / OBJECTIVES / HYPOTHESES

### 3.1. Aim

Phase 3 aims to characterise head injury presentations associated with cycling and e-scooter use at The Canberra Hospital and examine associations with helmet use and injury severity.

### 3.2 .Primary Objectives

- To quantify the number and characteristics of emergency department presentations for cycling- and e-scooter-related head injuries during the study period.
- To examine associations between helmet use and injury severity where helmet status is documented.

### 3.3. Secondary Objectives

- To describe injury patterns by age, sex, and vehicle type.
- To assess temporal trends in head injury presentations during the study period.

### 3.4. Hypotheses

- Compliance will vary by vehicle type, age, and gender presentation.
- Lower helmet use will be associated with higher rates of head injury presentations.

## 4. STUDY PHASE

- Phase 3: Retrospective Hospital Data Analysis

De-identified emergency department records from The Canberra Hospital will be retrospectively analysed to assess head injury presentations associated with cycling and e-scooter use during the study period. This component enables population-level linkage between observed helmet compliance trends and clinically significant outcomes, providing an objective measure of public health impact.

## 5. RESEARCH PLAN / STUDY DESIGN

### 5.1. Data Sources / Collection

#### 5.1.1. Retrospective Hospital Data

De-identified emergency department records from The Canberra Hospital will be extracted for cycling- and e-scooter-related head injuries occurring during the study period. Extracted variables will include age, sex, vehicle type, helmet use (if documented), and injury severity. This information will allow assessment of population-level associations between helmet compliance trends and clinically significant outcomes.

#### 5.1.2. Data Management and Security

All data will be de-identified and assigned unique codes for linkage and analysis. Hospital records will be stored on secure, password-protected institutional servers accessible only to authorised personnel. Raw data will not be shared externally. Data management procedures will comply with institutional, NHMRC, and local privacy guidelines, with routine backups and secure retention for a minimum of five years.

#### 5.1.3. Quality Control and Fidelity Monitoring

Coding will be conducted by trained researchers to ensure reliability. Regular review meetings will monitor data integrity, adherence to protocol, and timely resolution of any discrepancies.

### 5.2. Selection Criteria

#### Inclusion Criteria

- Emergency department presentations involving cycling or e-scooter use.
- Diagnoses indicating head injury, concussion, skull fracture, intracranial injury, or related trauma.
- Presentations occurring during the study period.

#### Exclusion Criteria

- Records lacking sufficient clinical detail to confirm the mechanism of injury.
- Duplicate records or follow-up visits for the same injury episode.

### 5.3. Statistical Analyses

All statistical analyses will be conducted in R®. Multiple imputation will address missing data where appropriate. Sensitivity analyses will include alternative specifications of period effects, adjustment for baseline covariates, per-protocol analyses limited to high-fidelity clusters, and subgroup analyses by age and baseline compliance.

#### 5.3.1. Hospital Data

De-identified emergency department records will be extracted by authorised ACT Health data custodians. Variables will include:

- Age
- Gender
- Vehicle type (e-bike, pedal bike, e-scooter, pedal scooter, where documented)
- Helmet use status (if recorded)
- Injury diagnosis and severity indicators (e.g., imaging results, admission status, ICU admission)
- Date and time of presentation

No names, addresses, medical record numbers, or other direct identifiers will be accessed by the research team.

### 6.4. Limitations

Incomplete emergency department documentation of helmet use is a known barrier experienced in previous studies (6). Consequently, helmet use may be subject to misclassification and missing data bias. Triage and emergency department staff will be informed of the study and reminded that helmet use may be documented when clinically relevant; however, no changes to routine clinical practice, documentation requirements, or triage processes will be requested.

## 6. ETHICAL CONSIDERATIONS

### 6.1. Recruitment and Selection of Participants

Hospital data will be accessed retrospectively under approved ethics protocols, with all records deidentified.

### 6.2 . Informed Consent

De-identified hospital data will be accessed under ethics approval; individual consent is not required.

### 6.3. Confidentiality and Privacy

All data will be de-identified and assigned unique codes. All data will be stored securely on encrypted servers accessible only to authorised personnel. Any external sharing for publication or collaboration will be fully anonymised and aggregated.

### 6.4. Data Access and Dissemination

Only study investigators and authorised data managers will access identifiable data (e.g., electronic medical records). Results will be disseminated at the aggregate level to policymakers, urban planners, cycling advocacy groups, and academic audiences.

### 6.5. Support for Staff

All staff will receive contact details for the research team and information on support services (e.g., Lifeline, Beyond Blue) if discussions of cycling injuries cause distress.

### 6.6. Aboriginal and Torres Strait Islander Data

The study does not seek to specifically identify Aboriginal or Torres Strait Islander children. Should the research team wish to analyse outcomes by Indigenous status, this will only occur with explicit consent and following approval from an Aboriginal Human Research Ethics Committee (HREC), in line with the NHMRC Ethical Conduct in Research with Aboriginal and Torres Strait Islander Peoples and communities (2018). The protocol will be amended and resubmitted for additional review if such analyses are proposed.

### 6.7. Data Storage and Record Retention

Electronic data will be stored on encrypted institutional servers, with routine backups maintained according to institutional and NHMRC guidelines. All data will be retained for a minimum of five years following study completion, after which secure deletion procedures will be implemented. Hard copies, if generated, will be securely shredded after the retention period. Data management plans will ensure compliance with applicable regulations, including the NSW Health Privacy Manual and NHMRC National Statement on Ethical Conduct in Human Research.

### 6.11. Ethics Approval and Oversight

Ethics approval will be sought from the University of Tasmania HREC and ACT Health Human Research Ethics Committee (HREC) before commencement of the relevant study phase. UTAS HREC will oversee all study phases, whilst ACTH HREC will also provide oversight in phase three. If Indigenous data collection is proposed, additional review will be sought from an Aboriginal HREC. All applications will be submitted and approved before any participant recruitment or data collection begins. The study protocol will be registered in the Australian and New Zealand Clinical Trials Registry (ANZCTR), and the registration details will be publicly available prior to study commencement. Publication of the protocol will also occur before the start of the study to ensure transparency and facilitate reproducibility. Continuous oversight of the study will be maintained throughout its duration, including monitoring participant safety, data integrity, and adherence to the approved protocol. Any adverse events will be promptly documented and reported to the HREC and relevant authorities, and participants will be informed of any developments that may impact their continued participation. Amendments to study procedures will be submitted for ethics review and approval before implementation.

## Data Availability

All data produced in the present study are available upon reasonable request to the author.

**LIST OF INVESTIGATORS AND PARTICIPATING INSTITUTIONS**

**Chief Investigator:**

Dr Brenton Systermans

Course Coordinator & Senior Lecturer, Healthcare in Remote and Extreme Environments,Tasmanian School of Medicine, University of Tasmania.

**Principal Investigator:**

Mr Alan Silburn

Paramedic & Registered Nurse (Division 2), NSW Ambulance / NSW Health

Academic, Western Sydney University

Fellow of the Royal Society for Public Health (FRSPH)

**Associate Investigator:**

Dr Sean Chan

Intensive Care Staff Specialist, The Canberra Hospital

State Medical Director, Donate Life ACT

Deputy Director, ACT Trauma Service

**Associate Investigator:**

Dr Thomas Georgeson

Emergency Staff Specialist, The Canberra Hospital

**Participating Institutions:**

University of Tasmania

Churchill Ave, Dynnyrne TAS 7005, Australia

The Canberra Hospital

Yamba Dr, Garran ACT 2605

## 7. OUTCOMES AND SIGNIFICANCE

The primary outcome of this study is to define the associations between observed helmet use trends and head injury presentations at The Canberra Hospital, offering a population-level perspective on the protective impact of helmets and the potential public health burden of non-compliance.

The significance of this study is reinforced when considered as part of the larger study. By integrating observational, clinical, and attitudinal data, it will generate a robust, context-specific evidence base for designing interventions aimed at improving helmet compliance among urban riders. Insights from this study may inform policy decisions, including the optimal deployment of educational signage, the calibration of deterrent fines, and broader urban planning strategies to enhance rider safety.

At a population level, the findings have the potential to reduce the incidence and severity of cycling and e-scooter-related head injuries, decrease healthcare utilisation, and promote a culture of safety in urban mobility. The evidence generated will be directly relevant to local policymakers, health authorities, transport planners, and advocacy organisations, supporting the development of scalable, cost-effective, and socially acceptable strategies to enhance helmet use. Ultimately, the study contributes to a broader understanding of behavioural determinants of safety compliance in rapidly evolving urban transport contexts and may serve as a model for similar initiatives in other Australian cities.

## 8. TIMELINES / MILESTONES

Months 0–2: Ethics approval, administrative access.

Months 2–4: Hospital data extraction.

Months 4–6: Data cleaning, coding, and analysis.

Months 6–12: Dissemination of results to stakeholders, manuscript preparation, and policy recommendations.

## 9. PUBLICATION POLICY

Findings will be disseminated via peer-reviewed publications, conference presentations, and reports to policymakers, urban planners, and advocacy organisations. Authorship will adhere to ICMJE guidelines.

## Declarations

### Abbreviations

Not applicable

### Human Ethics

Approval from the ACT Health Human Research Ethics Committee (HREC) will be obtained before Phase 3 commencement.

Ethics approval will then be obtained from the University of Tasmania Human Research Ethics Committee (HREC).

### Access

Approval to access the electronic medical records data at The Canberra Hospital will be requested before commencing Phase 3.

### Registration

The Helmet Use in Canberra study has been registered in the Australian and New Zealand Clinical Trials Registry (ANZCTR) [ACTRN12626000245392].

### Consent for publication

The author consents to the publication of this protocol.

### Availability of data and materials

Available at request from the corresponding author.

### Competing Interests

The author declares that they have no known competing interests or personal relationships that could have appeared to influence the work reported in this paper.

### Funding

The author declares that they did not receive funding for this article.

### Authors’ contributions

The author solely contributed to the conception, design, analysis, and drafting of the manuscript.

## Acknowledgements

Not applicable

## Notes

### Competing Interest Statement

The authors have declared no competing interest.

### Clinical Trial

ACTRN12626000245392

### Funding Statement

This study did not receive any funding.

### Author Declarations

Ethics approval will be sought from the University of Tasmania HREC and ACT Health Human Research Ethics Committee (HREC) before commencement of the relevant study phase. UTAS HREC will oversee all study phases, whilst ACTH HREC will also provide oversight in phase three.

